# Seroprevalence of Antibodies to SARS-CoV-2 in US Blood Donors

**DOI:** 10.1101/2020.09.17.20195131

**Authors:** Ralph R. Vassallo, Marjorie D. Bravo, Larry J. Dumont, Kelsey Hazegh, Hany Kamel

## Abstract

**Background:** To identify blood donors eligible to donate Coronavirus Disease-2019 (COVID-19) Convalescent Plasma (CCP), a large blood center began testing for antibodies to SARS-CoV-2, the etiologic agent of COVID-19. We report the seroprevalence of total immunoglobulin directed against the S1 spike protein of SARS-CoV-2 in US blood donors.

**Methods:** Unique non-CCP donor sera from June 1–July 31, 2020 were tested with the Ortho VITROS Anti-SARS-CoV-2 total immunoglobulin assay (positive: signal-to-cutoff (S/C) ≥1). Donor age, sex, race/ethnicity, ABO/RhD, education, and experience were compared to June and July 2019. Multivariate regressions were conducted to identify demographics associated with the presence of antibodies and with S/C values.

**Results:** Unique donors (n=252,882) showed an overall seroprevalence of 1.83% in June (1.37%) and July (2.26%), with the highest prevalence in northern New Jersey (7.3%). In a subset of donors with demographic information (n=189,565), higher odds of antibody reactivity were associated with non-Hispanic Native American/Alaskan (NH-NAA/A) and Black (NH-B), and Hispanic (H) race/ethnicity, age 18-64, middle school or lesser education, blood Group A, and never or non-recent donor status. In positive donors (n=2,831), antibody signal was associated with male sex, race/ethnicity (NH-NAA/A, NH-B and H) and geographic location.

**Conclusions:** Seroprevalence remains low in US blood donors but varies significantly by region. Temporal trends in reactivity may be used to gauge the effectiveness of public health measures. Before generalizing these data from healthy donors to the general population however, rates must be corrected for false positive test results among low prevalence test subjects and adjusted to match the wider demography.

## Introduction

Convalescent plasma is an investigational treatment for patients with coronavirus disease 2019 (COVID-19) that was recently granted Emergency Use Authorization (EUA).^1^ Most people develop neutralizing antibodies to its etiologic agent, Severe Acute Respiratory Syndrome Coronavirus-2 (SARS-CoV-2) during recovery from symptomatic illness. Historically, passive immune therapy has shown utility in other respiratory diseases (SARS-1, MERS, H1N1 and Spanish influenza).^2^ As such, antibody-containing COVID-19 convalescent plasma (CCP) from recovered donors has been transfused to those at risk for progressive disease in hopes of preventing life-threatening COVID-19. In a meta-regression analysis of seven small matched-control and randomized, controlled trials assessing the efficacy of CCP, a 57% odds reduction in mortality was observed in CCP recipients (absolute risk reduction 12%).^3^ In a study of >35,000 CCP recipients, significant reductions in 7- and 30-day mortality were correlated with earlier transfusion and higher immunoglobulin unit content.^4^ Accordingly, there is a need to identify and recruit individuals for CCP donation who have sufficient levels of circulating neutralizing antibodies against SARS-CoV-2 to meet rising demand and prepare a strategic reserve against successive waves of infection.

Recruiting individuals for CCP donation is challenging. Most programs began with hospitals referring recovered COVID-19 patients to blood collection establishments.^5^ The need for CCP has been publicized by governmental agencies, blood banking organizations, hospitals and blood collectors. Despite ample news outlet coverage, shortages of CCP occurred at program inception and again with case resurgence after relaxation of pandemic control restrictions. Eligibility criteria for CCP donation have evolved since the March 2020 FDA announcement of an emergency Investigational New Drug (IND) pathway for CCP administration. On April 3, 2020, FDA announced an expanded access program collecting open-label safety data, which just recently closed after FDA issued its EUA on August 23.^4,5^ To be effective, it is believed that CCP should contain adequate amounts of SARS-CoV-2 neutralizing antibodies. Since laborious tissue culture techniques are required for this assay, clinical criteria and binding antibody tests have been proposed as surrogates. Donation criteria have included allogeneic plasma-eligibility with evidence of COVID-19 documented by laboratory testing, with complete resolution of symptoms for at least 14 days and SARS-CoV-2 neutralizing antibody titers, if available. Throughout successive iterations of FDA Guidance, CCP donor eligibility criteria have remained the same.^5^

SARS-CoV-2 and other respiratory infections are not believed to represent a significant risk to blood safety.^6,7^ Screening blood donations for SARS-CoV-2 virus appears unwarranted. However, screening blood donors by serological methods to identify individuals with SARS-CoV-2 antibodies could serve other purposes. Surveillance studies document significantly higher numbers of infections than those defined by reported cases.^8^ This reflects individuals with asymptomatic illness, those with milder symptoms who did not seek medical care, and those who could not undergo testing. Because of the reduced positive predictive value of antibody testing in low prevalence groups, orthogonal testing and statistical adjustments must occur to assist in interpretation and generalize data to larger populations. Unadjusted serial sampling for SARS-CoV-2 antibodies can assess community viral spread in relation to containment measures or their relaxation. Antibody studies can also determine demographic risk factors, allow assessment of the durability of the immune response, and potentially detect reinfection when signal strength intensifies. Most important, high-throughput semi-quantitative binding antibody assays correlating with defined titers of neutralizing antibodies may identify and qualify donors for CCP donation while also identifying units with higher antibody titers.

Global efforts are ongoing to determine the country-specific prevalence of SARS-CoV-2 antibodies. However, in the vast majority, less than 10% of the population has been exposed to the virus, far below the 40-60% thought necessary for herd immunity.^9,10^ We describe the first two months’ results of serologic testing of healthy blood donors using a single total binding antibody assay conducted by the largest independent US blood center and its affiliate to help identify donors eligible to provide CCP.

Despite the best performance in a study of four high-throughput tests commonly available in the US, the S/C ratio in the Ortho VITROS Immunodiagnostic Products Anti-SARS-CoV-2 Total Ig test (αCoV2TIg), which we used to screen donors, does not precisely correlate with neutralization titers.^11,12^ S/C ratios on this platform do however, correlate with the presence of binding antibody across the test’s high dynamic range. Ratios have been reported to be higher in those recovering from more-severe COVID-19.13 We thus also sought to characterize the signal strength of positive samples.

## Methods

### Donor Testing

Vitalant began testing all allogeneic whole blood and apheresis donors for SARS-CoV-2 antibodies on June 1, 2020 to identify individuals potentially eligible to donate CCP. LifeStream, a Vitalant affiliate, commenced testing on June 8, 2020. Donors are asked to refrain from donating within 28 days of COVID-19 symptoms or diagnosis. They are informed about antibody testing and give consent for the research use of anonymized information, blood and blood samples. Test results are reported to donors using a secure portal for information and recruitment purposes.

Serum is tested using sample tubes collected with successful donations and routinely sent to Creative Testing Solutions (CTS - Bedford, TX; St. Louis, MO; Tampa, FL; Tempe, AZ). SARS-CoV-2 antibody testing uses the VITROS Immunodiagnostic Products Anti-SARS-CoV-2 Total Reagent Pack (αCoV2TIg, Ortho Clinical Diagnostics, Rochester, NY) on VITROS XT 7600 and 3600 instruments. The test has been FDA-authorized under an Emergency Use Authorization as an aid in identifying individuals with an adaptive immune response to the virus. αCoV2TIg is a chemiluminescent immunometric test qualitatively measuring total antibody (IgG, IgM and IgA) to SARS-CoV-2 S1 spike antigen. Results with signal-to-cutoff (S/C) ratios ≥1 are reported as positive.

CTS transmits test results and S/C ratios to centers’ blood establishment computer systems (BECS). A data warehouse extract (excluding CCP donations) was performed for all donations from June 1 to July 31, 2020 in our six BECS instances which included anonymized, encoded donation numbers and donor identifiers along with test results. Unique donor results were used to characterize the rate of COVID-19 antibody reactivity within geographic collection regions. Anonymized donor demographics were available for the largest BECS instance (eProgesa, MAK-SYSTEM, Paris, France) comprising ∼75% of tested collections. This subset database included:

- Donor demographics: age, sex, race/ethnicity, ABO/RhD, highest education level, and experience (first-time, active [prior presentation within 2 years], or reengaged [prior presentation >2 years ago])
- Donation information: date, Vitalant regional collection center, drive type (fixed site or mobile), and antibody test S/C result.

First-donation data were used if there was more than one tested donation in the analyzed time periods.

### Statistics

Donor demographic data for non-CCP donations from June 1 to July 31, 2020 were compared with the those from successful donations obtained on the same dates in 2019 for a subset of donors from the eProgesa BECS which contains information about donor race/ethnicity and highest education level. These proportions were compared using chi-square tests.

This subset database was also used to conduct an analysis of donor demographics associated with the presence of antibodies and with S/C ratios. An unadjusted logistic regression was initially performed to assess the crude association of each category with a positive antibody test. Multivariate logistic analysis was then performed to calculate the adjusted odds ratios (OR) and 95% confidence intervals (CIs) (Stata 15.1, Stata Corp., LLC, College Station, TX, USA). The final model included all donor and donation characteristics. The log10-transformed S/C from donors with positive αCoV2TIg test results were regressed against seven donor demographic characteristics and blood type using general estimating equations for main effects (Proc Mixed, SAS v9.4, SAS Institute, Cary, NC). Main effects with p<0.05 were retained in the final model. The log10 S/C ± 2SE was estimated from the final model, then converted back to linear S/C (estimated geometric mean ± 2SE). Estimated differences between categories were assessed using the model.

## Results

From June 1 through July 31, 2020, 254,499 unique donors completed a whole blood or non-CCP apheresis donation of which 99.4% yielded tubes tested for SARS-CoV-2 antibodies (Table 1). Figure 1 shows the changing rate of antibody reactivity across the geographies in which Vitalant and its affiliate collect blood. There was a demonstrable increase in overall seroprevalence from 1.37% in June to 2.26% in July. Of the 16,383 individuals donating more than once during this time period, 35 donors (0.2%) seroconverted between their first and a later donation within the 2-month time period.

**Table 1.** Donation and Unique Donor Totals for June/July 2019 and 2020 Across all Vitalant/LifeStream Centers and within the Subset with Expanded Demographics (combined number of unique donors is smaller than the sum of both months due to some donors’ return in the second month)

|  | **All** | **Demographics Subset** |  |
| --- | --- | --- | --- |
|  | **June / July 2020** | **June / July 2019** | **June / July 2020** |
| **Successful Phlebotomies** | 140,617 / 130,510 | 88,231 / 87,281 | 105,859 / 97,586 |
| **Unique Donors** | 137,041 / 126,869<br>(254,499 both months) | 85,322 / 84,078<br>(162,394 both months) | 103,121 / 94,769<br>(190,595 both months) |
| **Donations Tested for Ab** | 138,790 / 130,475 |  | 105,462 / 97,012 |
| **Unique Donors** | 135,353 / 126,808<br>(252,882 both months) |  | 102,732 / 94,202<br>(189,656 both months) |

**Figure 1.**
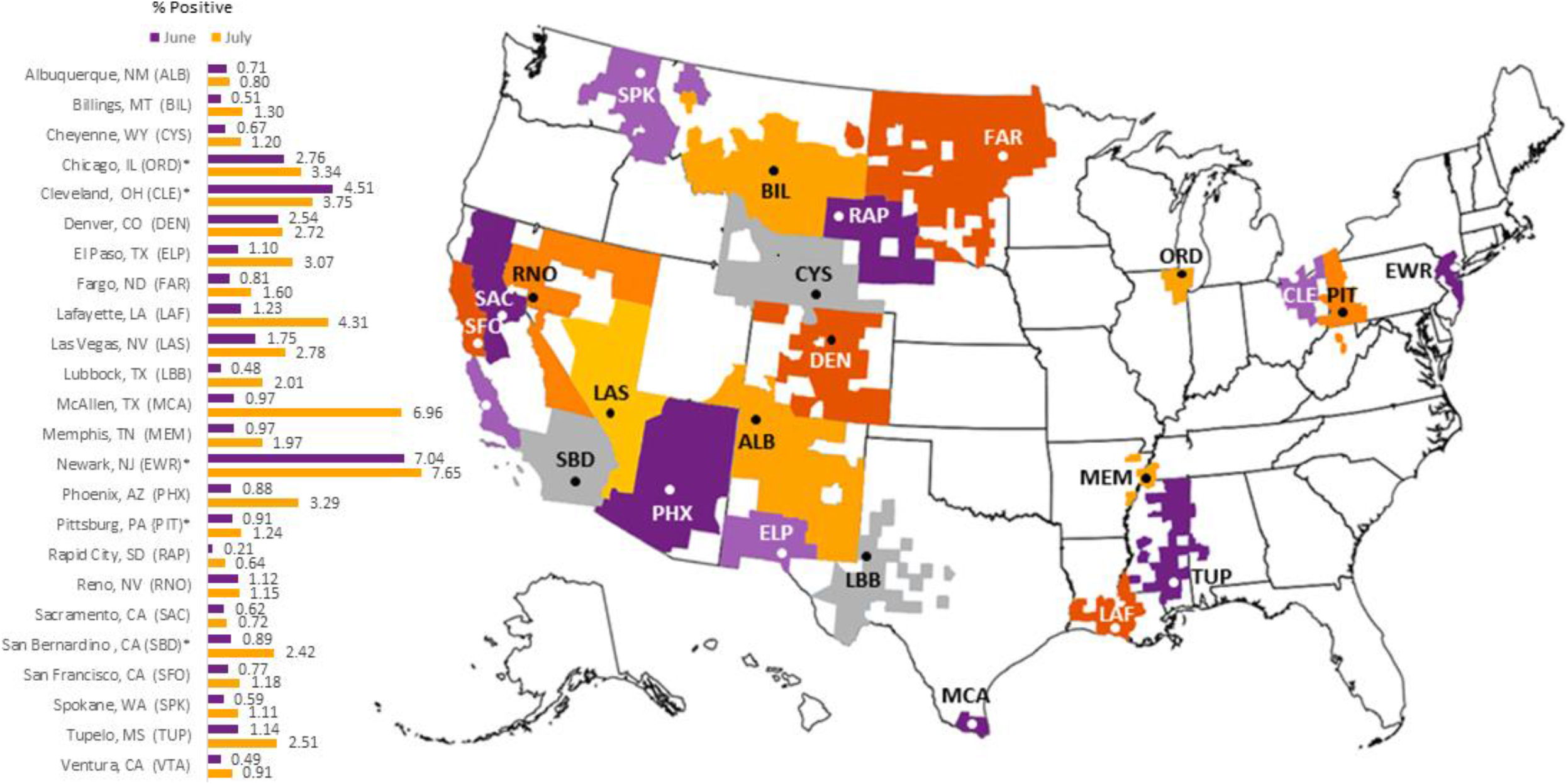
Vitalant/LifeStream Regional Center Collection Areas and % Positive for SARS-CoV-2 Antibodies

Comparison data from the large subset of donors for which expanded demographic information was available revealed significant differences in donor composition between the same time periods in 2019 and 2020 (Table 2), in addition to greater overall turnout (Table 1). COVID-19 public gathering restrictions resulted in a 19% reduction in mobile collections, while fixed site donations rose by 59%. Of individuals successfully donating, increases were seen in the fraction of females, non-Hispanic Whites, those with post-high school degrees and people aged 30 and over. Greater numbers of first-time and reengaged donors also chose to donate. Unpublished data from April and May 2020, before donor testing was offered, showed demographic shifts similar to those observed in June and July 2020.

**Table 2.**
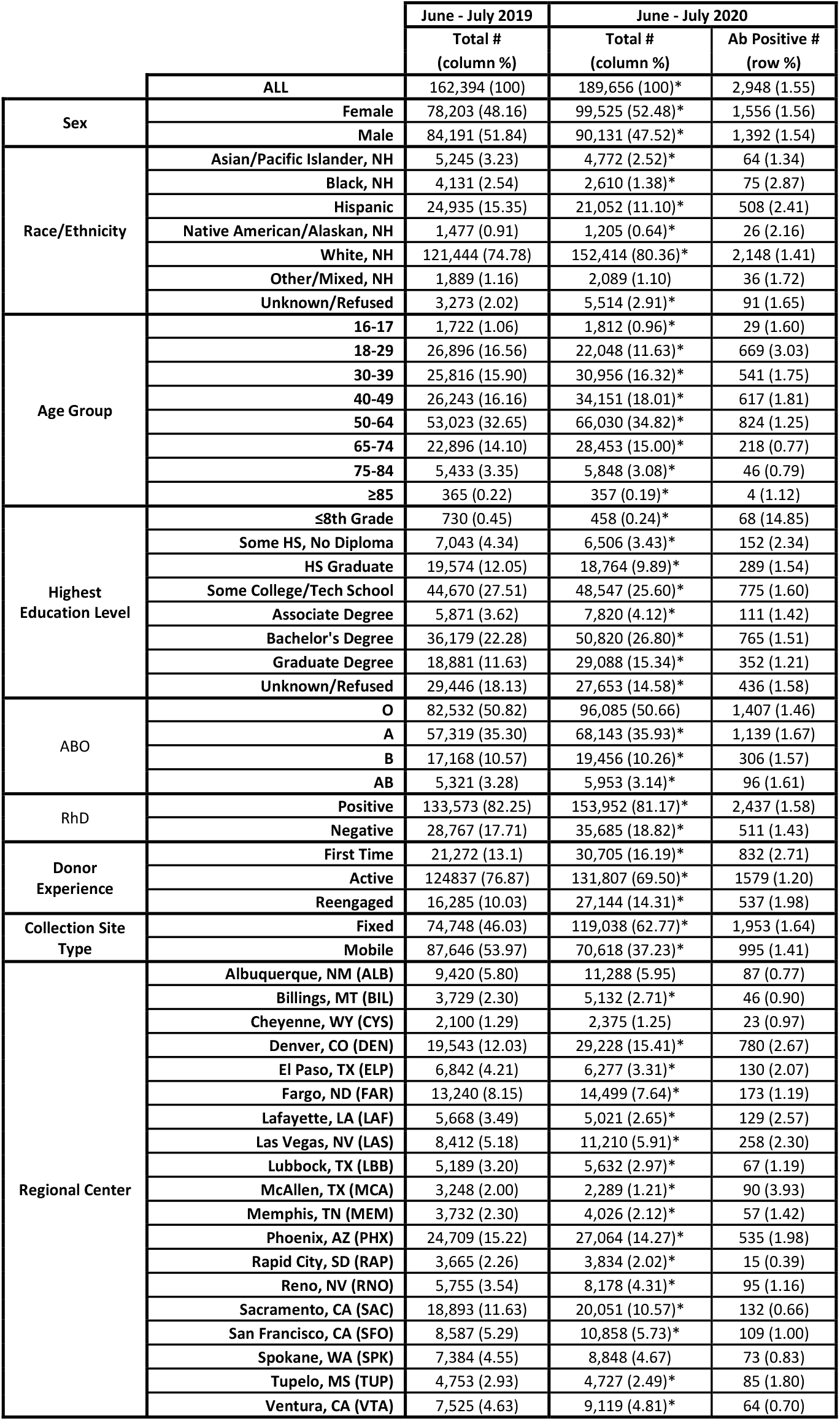
Donor Demographics and Donation Information in Subset Centers, June/July 2019 and 2020 (* denotes univariate comparisons that are statistically-significantly different [p<0.05] between years)

Univariate comparisons of antibody positivity by sex, race/ethnicity, age, education, blood group, donor experience, collection site type and region are presented in Table 2. Results of the multivariate analysis for factors associated with positive antibody test results by donor and donation characteristics is presented in Figure 2. While no significant differences were observed for sex and RhD, there were differences by age, race/ethnicity, education, ABO blood group, donation experience, collection site type and region. Compared with non-Hispanic White donors, those who are non-Hispanic Native Americans/Alaskans and Blacks, or Hispanic donors were significantly more likely to have a positive test result. Individuals aged 18-29 had the highest odds of a positive result, but those 30-64 were also significantly more likely than those under 18 or over 64 to be positive. Compared with individuals with a high-school degree, those with no high-school experience were much more likely to test positive. Donors with graduate degrees were slightly less likely to have a positive test result. A small but statistically-significant increase was seen in blood Group A donors compared with Group O individuals. First-time and reengaged donors also had a higher risk for antibody-positive results. Donors at the reduced number of mobile drives had a slightly lower risk for antibody positivity. Figure 2B shows the adjusted geographical differences in seroprevalence, expected on the basis of reported variations in community COVID-19 incidence over time.

**Figure 2.**
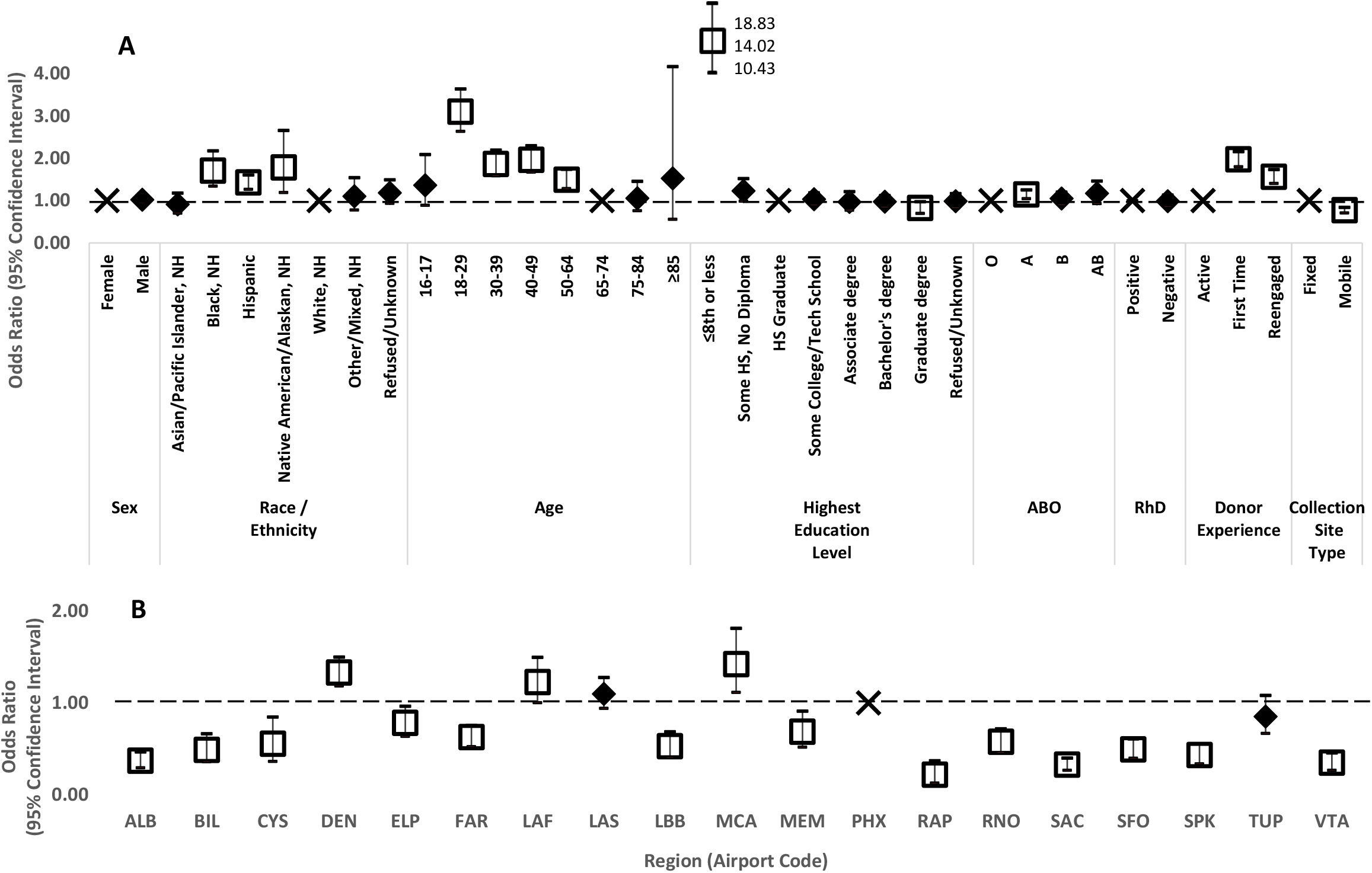
Multivariate Analysis of Predictors of COVID Antibody Positivity for 189,656 Unique Donors, June/July 2020; A: Donor Demographics; B: Donation Location. (**X**: reference category, **□**: OR (95% CI) statistically-significantly different from reference; **♦**: OR (95% CI) not significantly different from reference)

The distribution of donor S/C values is shown in Figure 3. In June and July 2020, 2,948 reactive donors had a αCoV2TIg S/C ≥ 1. The first donor visit S/C was available for analysis for 2,831 donors (median 68.8, IQR 17.0-177, geometric mean 48.2, maximum 961). Log10 transformation demonstrates a left-skewed distribution (Figure 3B). Supplemental Table 1 outlines the log10 and back-transformed S/C data in 2,831 donors with positive antibody test results.

**Figure 3.**
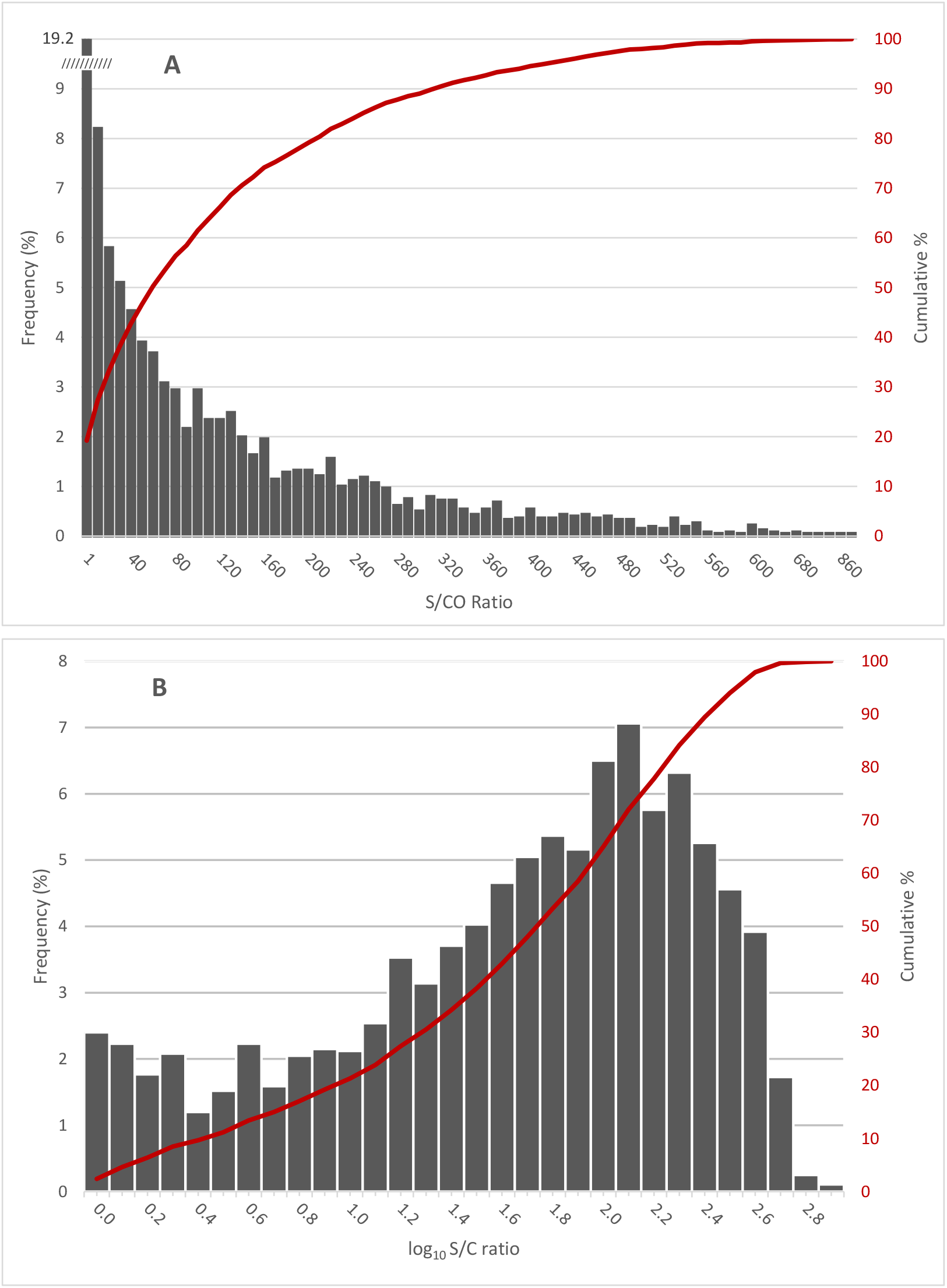
Distribution of S/C Ratios for Positive Samples (S/C ≥1.0). A: Linear Frequency with Cumulative Percentages; B: Log10 S/C Distribution with Cumulative Percentages

We further examined the association of donor and donation site characteristics with S/C in αCoV2TIg-positive donors. The association of 7 donor demographic characteristics and blood type was explored with the log10-transformed S/C in a multivariate regression model of these main effects (Table 3). Highest education level, collection site type, and ABO-RhD interaction were found to have insignificant contributions and were removed from the model. Main independent variables with significant effects on the S/C were donor sex, race/ethnicity, donor experience, and regional center. Variable subcategories are shown in Figure 4 and the fully-adjusted model-estimated S/C ± 2SE compared to the overall model-estimated geometric mean (45.2). Females had small, but significantly lower S/Cs than males; non-Hispanic Native American/Alaskan, non-Hispanic Black, and Hispanic groups’ values were significantly higher than the average; and the Denver Region had the highest adjusted S/C of all 19 examined. The overall effect of donor experience was significant. Although not significantly different from the model mean (p>0.05), first-time donors’ S/Cs are an estimated 1.3 units higher than active donors’ (p=0.0004).

**Table 3.**
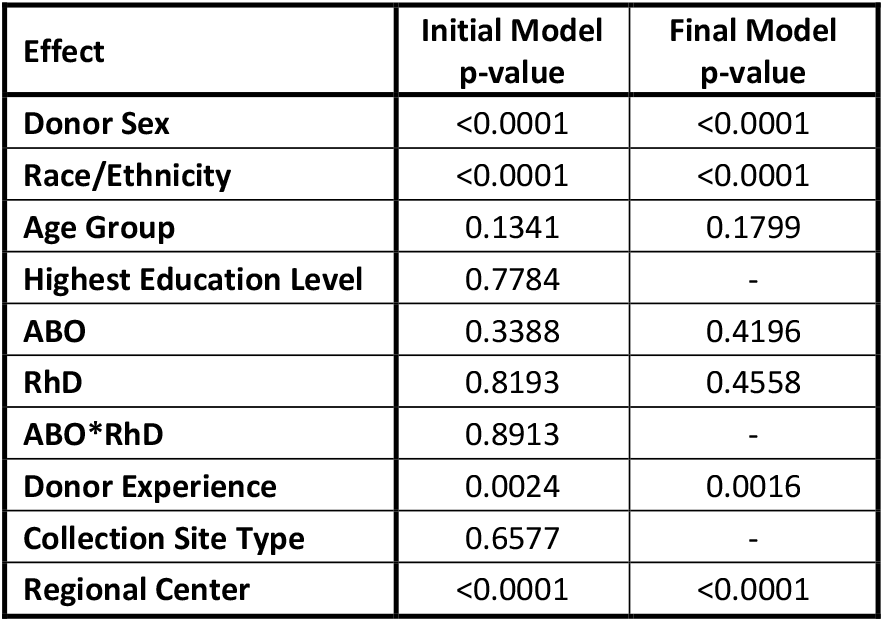
Positive-Antibody Donor Main Effects Model

| Effect | Initial Model<br>p-value | Final Model<br>p-value |
| --- | --- | --- |
| Donor Sex | <0.0001 | <0.0001 |
| Race/Ethnicity | <0.0001 | <0.0001 |
| Age Group | 0.1341 | 0.1799 |
| Highest Education Level | 0.7784 | - |
| ABO | 0.3388 | 0.4196 |
| RhD | 0.8193 | 0.4558 |
| ABO*RhD | 0.8913 | - |
| Donor Experience | 0.0024 | 0.0016 |
| Collection Site Type | 0.6577 | - |
| Regional Center | <0.0001 | <0.0001 |

**Figure 4:**
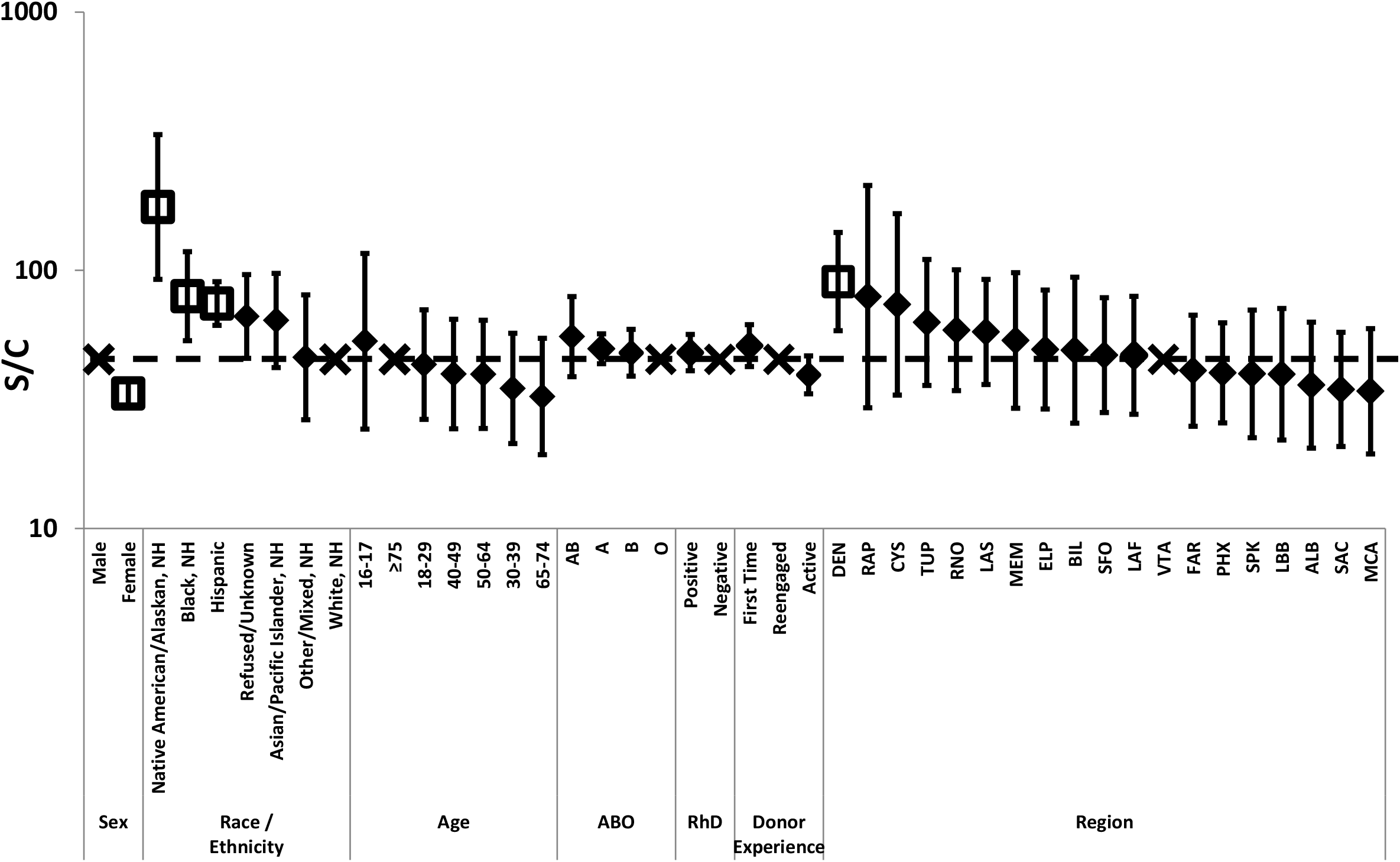
Positive-Antibody Donor S/C Multivariate Model Estimates – The classification variables of donor sex, race/ethnicity, and collection region had a significant effect on the S/C estimate (p<0.015). Shown are estimated geometric means [± 2SE] for subcategories within classification variables. Estimated overall geometric mean was 45.2 [23.5-86.6]. The overall effect of donor experience was significant. Although not significantly different from the model mean (p>0.05), first-time donors’ S/Cs are an estimated 1.3 units higher than active donors’ (p=0.0004). (**X**: reference category, **□**: estimate statistically-significantly different from reference; **♦**: estimate NOT significantly different from reference)

## Discussion

We present seroprevalence data from a large cohort of geographically-diverse healthy blood donors over the first two months of SARS-CoV-2 antibody testing on the αCoV2TIg platform. The overall seropositive rate was 1.83% amongst 252,882 unique donors tested. The rate in the second month of testing was higher than the first month. In multivariable analysis, higher reactivity rates were associated with middle school or lesser education (unadjusted point estimate, 14.8% positive), age (highest result [3.0%] in those 18 to 29 years old), race/ethnicity (highest point estimate in non-Hispanic Blacks [2.9%]), and first-time donors (2.7% compared with 1.2% in active donors). There was a small, but statistically-significant increase in reactivity seen in blood Group A donors, and a lower risk in donors attending the declining number of mobile collections. Significant regional differences were observed, with June/July unadjusted positive rates highest at 7.3% in the Newark, NJ area and the lowest rate (0.4%) south/east of Rapid City, SD.

SARS-CoV-2 serosurveillance studies are beginning to appear in the literature. As data are compared, it is important to consider the characteristics of the population studied, geographic location, dates of testing, and screening laboratory methods. A 13.7% prevalence of SARS-CoV-2 antibodies was reported in 40,329 healthcare personnel in the New York City area, tested from April to June 2020.^14^ This study employed 7 different assays of spike- and nucleocapsid-directed IgG and total antibody. Two relatively small reports in Canadian blood donors using the Abbott Architect IgG assay revealed low rates: 2.23% of 7,691 donors in Quebec tested over a 6-week period and <1% of 10,000 blood donors in one month of testing from all other provinces.^15^

Demographic associations of test positivity in blood donors have not yet been reported. Compared with a similar period in the preceding year, the demography of blood donors significantly changed with the enactment of COVID-19 safety requirements. Contributors to shifting donor demographics included the replacement of cancelled mobile drives with more-limited geography fixed site appointments, ongoing needs messaging via public service announcements and perhaps, the announcement of testing in June. This observed demographic shift was similar to other, shorter-lived post-disaster responses.^16^ Donors responding to messaging about the need for blood increased the year-over-year number of summer donations. Female donors, non-Hispanic Whites, those with post-high school degrees and donors over age 29 disproportionately responded to appeals. A greater proportion of first-time and former blood donors answered the call. As noted, this was not clearly linked with the initiation of SARS-CoV-2 antibody testing as these demographic shifts were present in April and May 2020 before CCP-eligible donor screening began. Greater fractions of antibody reactivity in new and reengaged donors does however, suggest some effect from the announcement of antibody testing.

SARS-CoV-2 binding Total Ig antibodies appear to be stable over months for individuals with previous infection and are thus suited for surveillance studies.^11,17^ While well suited for surveillance, these results do not correlate as closely with neutralizing antibody titer, which may be an important direct or proxy measure of CCP potency. Other assays in which antibody reactivity wanes over time are less suited to detect previously-infected individuals but tend to correlate more closely with neutralizing antibody titer.^18^ In our study, higher S/C values for spike Total Ig were associated with male sex, non-Hispanic Native American/Alaskan and Black race/ethnicity, and Hispanic ethnicity. Whether this correlates with the severity of infection or indicates a more vigorous immunologic response remains to be determined. First-time donors and individuals in geographies with a higher prevalence of antibody reactivity also had higher S/C values.

FDA recently defined a “high” neutralizing antibody titer cutoff in a specific assay associated with a 21% reduction in 7-day mortality amongst non-intubated patients compared with recipients of lower-titer CCP units in the Mayo expanded access program.^19^ FDA’s Center for Biologics Evaluation and Research (CBER) has stated its intention to review the acceptability of alternative binding antibody tests to be used to designate high-versus low-titer CCP, which must be labeled as such beginning December 1, 2020.^20^ Identifying donors more likely to have higher antibody titers is thus a meaningful exercise.

This study has several limitations. First, this is an initial report on the first two months of screening blood donors for antibodies to SARS-CoV-2; continued reporting is necessary to define trends as public health measures are enacted and relaxed. Second, there is a lack of donor clinical information (symptomatology and laboratory confirmation of infection by PCR) and no orthogonal supplemental testing has been conducted to weed out false-positive test results in this low-prevalence population. These limit the ability to correlate antibody reactivity and S/C strength with the presence or severity of symptoms. Last, blood donors are a significantly healthier population than the general public, with substantially different demography than the general population. Generalization of data from this group of tested individuals to the population at large must be corrected for potential false positive test results and adjusted to reflect the demography of the general population before estimates of pathogen exposure are valid. Trends over time however, are valuable in the evaluation of imposition or relaxation of efforts to limit spread of the disease. While longitudinal studies are under way to better characterize pathogen exposure, data like ours also provide a rapid estimate of regional differences in the number of infected individuals as we progress through substantial morbidity and mortality toward herd immunity.^10^

As commonly seen in the setting of traumatic events, blood donor demographics changed during the COVID-19 pandemic. We reported on the prevalence of SARS-CoV-2 antibodies and the correlates influencing reactivity. Our report complements the growing number of surveillance studies in various populational groups.

## Data Availability

All data referred to in the paper are contained within its tables and figures.

## Acknowledgement

The authors would like to acknowledge the contributions of D. Joseph Chaffin, MD and Frederick Axelrod, MD of LifeStream.

---

**Supplemental Table 1.**
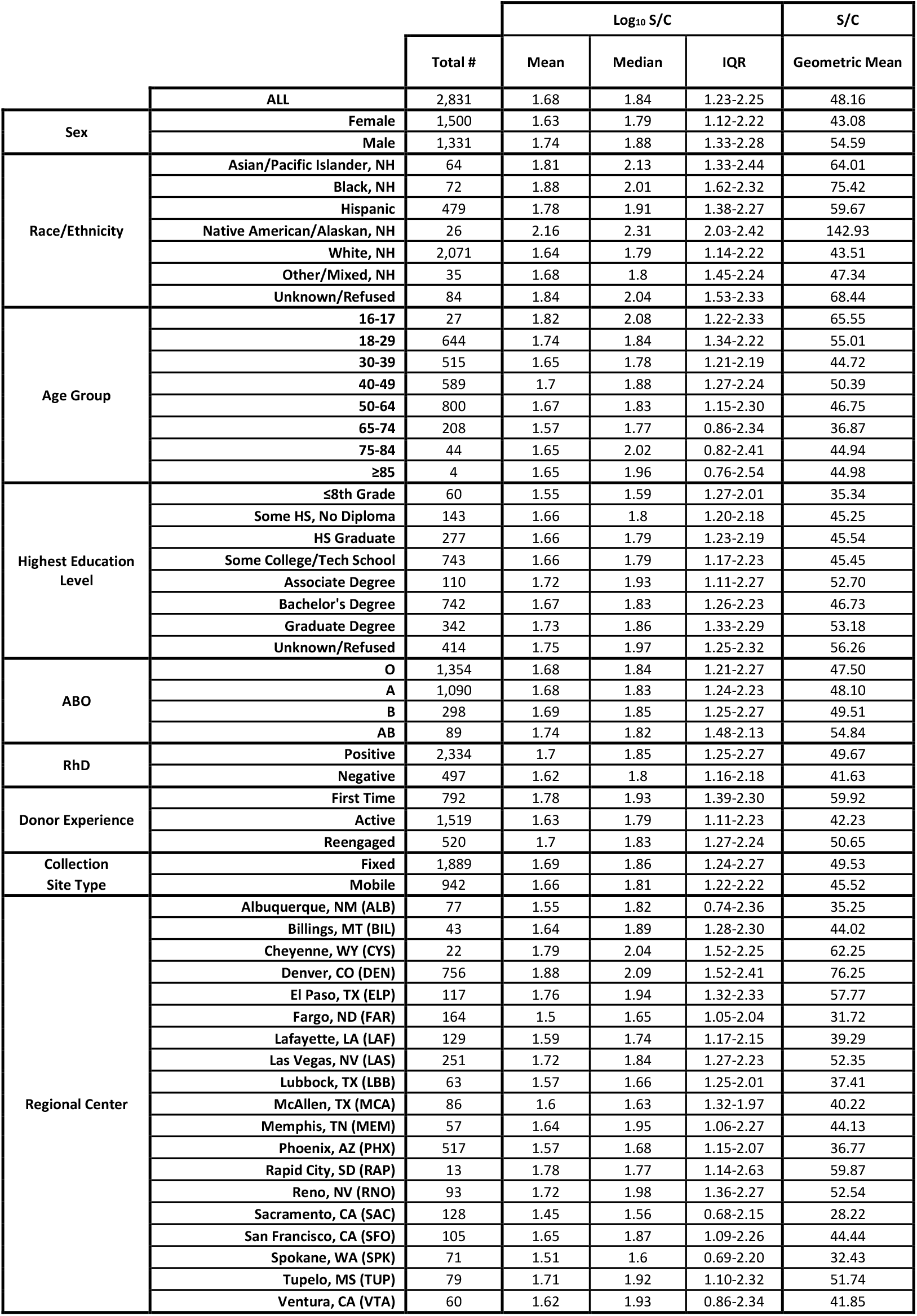
Summary Statistics of Signal-to-Cutoff (S/C) Ratios

## Notes

**SOURCES OF SUPPORT**: Blood Center Foundation of the Inland Northwest, Blood Science Foundation, Bonfils Blood Center Foundation Donor-Advised Fund, CDC Contract 75D301-20-C-08170, Vitalant

**CONFLICTS OF INTEREST**: Drs. Bravo, Dumont, Hazegh, Kamel and Vassallo have declared no relevant potential conflicts of interest.

### Competing Interest Statement

The authors have declared no competing interest.

### Funding Statement

Support for SARS-CoV-2 Antibody testing was provided by Vitalant and its contributors: Blood Center Foundation of the Inland Northwest, Blood Science Foundation, Bonfils Blood Center Foundation Donor-Advised Fund. Additional funding for data-sharing was provided under CDC Contract 75D301-20-C-08170.

### Author Declarations

Advarra has certified this research effort exempt (Protocol 00030878).

## References

1. Hinton DM. August 23, 2020 letter to the Assistant Secretary for Preparedness and Response authorizing the emergency use of COVID-19 convalescent plasma for treatment of hospitalized patients with COVID-19. Available at: https://www.fda.gov/media/141477/download. Last accessed September 10, 2020.

2. Mair-Jenkins J, Saavedra-Campos M, Baillie JL, et al., for the Convalescent Plasma Study Group. The effectiveness of convalescent plasma and hyperimmune immunoglobulin for the treatment of severe acute respiratory infections of viral etiology: a systematic review and exploratory meta-analysis. J Infect Dis 2015;211:80–90.

3. Joyner MJ, Klassen SA, Senefeld JW, et al. Evidence favouring the efficacy of convalescent plasma for COVID-19 therapy. MedRxiv 2020;Jul 30. doi: 10.1101/2020.07.29.20162917.

4. Joyner MJ, Senefeld JW, Klassen SA, et al., for the US EAP COVID-19 Plasma Consortium. Effect of convalescent plasma on mortality among hospitalized patients with COVID-19: initial three-month experience. MedRxiv 2020;Aug 12. doi: 10.1101/2020.08.12.20169359.

5. Budhai A, Wu AA, Hall L, et al. How did we rapidly implement a convalescent plasma program? Transfusion 2020;60:1348–55.

6. Katz LM. Is SARS-CoV-2 transfusion transmitted? Transfusion 202060:1111–4.

7. Andersson M, Arancibia-Carcamo CV, Auckland K, et al. SARS-CoV-2 RNA detected in blood samples from patients with COVID-19 is not associated with infectious virus. MedRxiv 2020;Jun 17. doi: 10.1101/2020.05.21.20105486.

8. Havers FP, Reed C, Lim T, et al. Seroprevalence of Antibodies to SARS-CoV-2 in 10 Sites in the United States, March 23-May 12, 2020. JAMA Intern Med 2020 Jul 21;ePub. doi: 10.1001/jamainternmed.2020.4130.

9. Arora RK, Joseph A, Van Wyk J, et al. SeroTracker: a global SARS-CoV-2 seroprevalence dashboard. Lancet Infect Dis 2020;S1473-3099(20)30631-9.

10. Britton T, Ball F, Trapman P. A mathematical model reveals the influence of population heterogeneity on herd immunity to SARS-CoV-2. Science 2020;369:846–9.

11. Padoan A, Bonfante F, Pagliari M, et al. Analytical and clinical performances of five immunoassays for the detection of SARS-CoV-2 antibodies in comparison with neutralization activity. MedRxiv 2020;Jun 17. doi: 10.1101/2020.08.01.20166546.

12. Goodhue Meyer E, Simmons G, Grebe E, et al. Selecting COVID-19 Convalescent Plasma for Neutralizing Antibody Potency Using a High-capacity SARS-CoV-2 Antibody Assay. MedRxiv 2020; Sept 2. doi: 10.1101/2020.08.31.20184895.

13. Nagura-Ikeda M, Imai K, Kubota K, et al. Clinical characteristics and antibody response to SARS-CoV-2 spike 1 protein using the VITROS Anti-SARS-CoV-2 antibody tests in COVID-19 patients in Japan. MedRxiv 2020;Aug 4. doi: 10.1101/2020.08.02.20166256.

14. Moscola J, Sembajwe G, Jarrett M, Farber B. Prevalence of SARS-CoV-2 antibodies in health care personnel in the New York City area. JAMA 2020 Aug 6;e2014765.

15. COVID-19 Community Taskforce. The COVID-19 immunity task force welcomes the results of the blood donor seroprevalence study led by Héma-Québec. Available at: https://www.covid19immunitytaskforce.ca/the-covid-19-immunity-task-force-welcomes-the-results-of-the-blood-donor-seroprevalence-study-led-by-hema-quebec/ Last accessed September 10, 2020.

16. Glynn SA, Busch MP, Schreiber GB, et al., for the NHLBI REDS Study Group. Effect of a national disaster on blood supply and safety: the September 11 experience. JAMA 2003;289:2246–53.

17. Gudbjartsson DF, Norddahl GL, Melsted P, et al. Humoral immune response to SARS-CoV-2 in Iceland. N Engl J Med. doi: 10.1056/NEJMoa2026116.

18. Muecksch F, Wise H, Batchelor B, et al. Longitudinal analysis of clinical serology assay performance and neutralising antibody levels in COVID19 convalescents. MedRxiv 2020;Aug 6. doi: 10.1101/2020.08.05.20169128

19. Redacted. EUA 26382: Emergency Use Authorization (EUA) Request Clinical Memorandum. Available at: https://www.fda.gov/media/141480/download. Last accessed September 10, 2020.

20. US Food and Drug Administration Center for Biologics Evaluation and Research. Investigational COVID-19 Convalescent Plasma: Guidance for Industry. Silver Spring, MD: September 2, 2020. Available at: https://www.fda.gov/media/136798/download. Last accessed September 10, 2020.

